# Reduced penetrance of *COL1A1/2* pathogenic variants linked with osteogenesis imperfecta: analysis of a large population cohort

**DOI:** 10.1101/2025.10.22.25338523

**Authors:** Alistair T. Pagnamenta, James Fasham, Robin N. Beaumont, Duncan Baker, Tim Hall, Emma L. Baple, Meena Balasubramanian, Leigh Jackson, Caroline F. Wright

## Abstract

Osteogenesis imperfecta (OI) is under consideration for inclusion in several genomic newborn screening initiatives, but its penetrance in clinically-unselected populations is currently unknown. It is an exemplar condition for evaluating penetrance in adult cohorts due to its relatively low mortality, variable expressivity and link to several large genes. Using genome sequencing data from ∼500,000 adults in UK Biobank, we curated a set of rare pathogenic/likely pathogenic variants in *COL1A1, COL1A2* and *IFITM5* using annotations from gnomAD, ClinVar and SpliceAI. Analysis of summed read-count data from genome and exome sequencing led to exclusion of 16 mosaic variants with consistently low allelic balance of 3.4-35.9%. We identified 61 likely constitutive heterozygous pathogenic variants in *COL1A1* and *COL1A2* (29 loss-of-function or splice variants and 32 missense) in 115 participants, with a mean age at recruitment of 55.1 years; no pathogenic variants were identified in *IFITM5*. Phenotypes were assessed using ICD-10 codes, self-reports and heel bone mineral density. Overall disease penetrance was lower than anticipated: 40.7% for *COL1A1* and 21.3% for *COL1A2*, potentially due to depletion of severe early-onset disease. When considering only truncating variants in *COL1A1*, disease penetrance increased to 73.1%, and 90% of individuals had reduced levels of circulating COL1A1 protein. For *COL1A2*, bone mineral density data supported the low penetrance, whereas for *COL1A1*, data suggested the possibility of a subclinical phenotype. Overall, for pathogenic missense variants (including those altering Gly-Xaa-Yaa repeats), the low penetrance observed suggests reliance on current ClinVar assertions to support pathogenicity may overstate OI risk in population screening.

## Introduction

Newborn screening (NBS) involves the widespread testing for specific genetic or metabolic disorders to enable early detection/diagnosis and facilitate improved clinical management. A well-defined set of criteria, based on the Wilson and Jungner principles (1968)^1^ are used to decide which conditions should be considered for inclusion in NBS programmes. For genetic conditions, screening should be limited to severe, early-onset diseases with high penetrance, i.e. where a pathogenic variant has a high likelihood (>80-90%) of causing the associated disease.^2^ Unfortunately, penetrance estimates in the scientific literature are typically based on clinical cohorts and may suffer from ascertainment bias.^3^ A recent study integrating electronic health records with predicted loss-of-function (pLOF) variants across 91 genes confirmed to cause disease via genetic haploinsufficiency indicates that incomplete penetrance is not uncommon.^4^ Therefore, to avoid over-reporting, it is imperative that the penetrance of all genetic conditions included in NBS initiatives be systematically re-evaluated using large population cohorts.

Over 30 countries are currently assessing the potential benefits of using genome sequencing to expand rapid NBS testing to include a larger number of genes/variants in parallel.^5^ One such initiative is the UK’s Generation Study,^6^ which commenced active recruitment in 2024 and currently reports pathogenic and likely pathogenic variants in >450 genes linked to >200 conditions. Despite extensive guidance on how gene-condition pairs may qualify for inclusion in NBS testing, there remains little consensus. For instance, comparison of 27 international genomic NBS studies investigated >4000 genes that were included in at least one study but highlighted that just 18 genes were included in all 27 genomic NBS initiatives.^5^

Osteogenesis imperfecta (OI) is a genetically heterogeneous bone dysplasia, characterized by low bone mass and increased bone fragility.^7,8^ Affected individuals are at increased risk of long bone fractures and vertebral compressions. Variable deformities of the long bones, ribs and spine may be observed in some cases from birth and there may also be substantial growth deficiency. Other features can include blue sclerae, hearing loss and dental anomalies such as dentinogenesis imperfecta. Studies from the US and across Europe have estimated the prevalence of OI at between 0.3 and 1.5 cases per 10,000 individuals.^7,9^ Of the 33 genes conclusively linked to OI (i.e. green on the OI panel in Genomic England’s PanelApp^10,11^, included in the NHS Genomic Medicine Service Test Directory and also included in PanelApp Australia’s OI and Osteoporosis panel), only three are currently in the inclusion list for the Generation Study. These include *COL1A1* (MIM: 120150) and *COL1A2* (MIM: 120160), which together represent the most common cause of OI (types I-IV),^7,9,12^ and *IFITM5* (MIM: 614757), which is linked to a rarer form of severe OI (type V) caused by a much smaller repertoire of activating variants.^13^

*COL1A1* and *COL1A2* are closely related paralogous genes that share a highly similar structure and encode the α1 and α2 collagen chains. These chains form a heterotrimeric triple-helix, composed of two α1 and one α2 molecules. Due to the tightly coiled nature of this structure (PDB:1CAG, 1.85Å),^14^ every third amino acid residue in the helical domains has evolved to be a glycine. Variants that disrupt this Gly-Xaa-Yaa pattern can severely disrupt the triple-helical structure and often result in acute phenotypic presentations, through a dominant-negative mechanism.^7^ Of the four distinct subtypes of OI caused by *COL1A1* and *COL1A2* variants, types II (a perinatal lethal form with death mostly due to respiratory insufficiency) and III (a ‘progressive deforming’ non-lethal form) are the most severe.^15^ In contrast, the milder OI type I is more often caused by pLOF variants in *COL1A1* that are predicted to reduce protein levels.^12^ Despite several genotype-phenotype studies, predicting clinical outcome is not straightforward, particularly for previously unreported variants. Studies based on clinical cohorts have suggested that although expressivity varies widely, even within families, disease penetrance in individuals carrying a heterozygous pathogenic variant in *COL1A1* or *COL1A2* is close to 100%.^8^

Management of newborns with a suspected genetic diagnosis of OI involves careful handling of the infant from birth, with avoidance of manual hip examination. Treatment may also include the use of bisphosphonates, surgical or physiotherapy interventions and ventilatory support, depending on clinical phenotype. Early adoption of such interventions, following a molecular confirmation of OI, have been associated with reductions in early morbidity and mortality.^9^ However, the potential benefits of such therapies must be weighed against the potential harms caused by the reporting of false positive results, which can include side-effects of unnecessary treatment, psychological distress for families and more generally, the loss of public trust in genetic testing. In addition, given the variable expressivity of OI, the value of early genetic testing is an area of debate, with the key benefit being advice on handling in newborns and infants. Conversely, some of these children may never present with a fracture in their lifetime and therefore implications around what appropriate follow up should be remain unclear. This also brings into question resources required to take on this additional workload for clinical teams. In this study, we evaluated the prevalence and penetrance of pathogenic and likely pathogenic variants in *COL1A1, COL1A2* and *IFITM5* in a population cohort of half a million UK adults. By integrating these variants with a rich set of clinical, molecular and physiological phenotypes, we assessed gene-level population penetrance in order to evaluate the efficacy of including these three genes in genomic NBS.

## Materials and Methods

### Genome sequencing and variant annotation

UK Biobank (UKB) is a population-based study involving half a million participants from across the UK, recruited between 2006-2010 at age 40–69 years.^16^ Data access was obtained under the approved UKB project ‘Understanding the role of rare and common genetic variation in human phenotypes’, application number 103356. In addition to the collection of comprehensive demographic and health-related measures, both exome (N=454,787) and genome sequencing (N=490,640) were performed for the majority of participants.^17,18^ In this study, variant curation primarily focussed on the genome sequencing data, which used 150bp paired-end reads and the NovaSeq 6000 sequencing instrument (Illumina). Mapping to GRCh38 and variant calling used the DRAGEN pipeline. Variants were annotated using Ensembl VEP (v110)^19^, as described elsewhere (Web resources) and SNVs/indels in *COL1A1, COL1A2* and *IFITM5* were extracted based on the MANE Select transcripts (ENST00000225964.10 / NM_000088.4 for *COL1A1*, ENST00000297268.11 / NM_000089.4 for *COL1A2* and ENST00000382614.2 */* NM_001025295.3 for *IFITM5*).

### Filtering and QC of variants

Different variant filtering approaches were taken for each of the three candidate genes, and a summary of prioritisation steps is provided below and in **Figure 1**.

**Figure 1:**
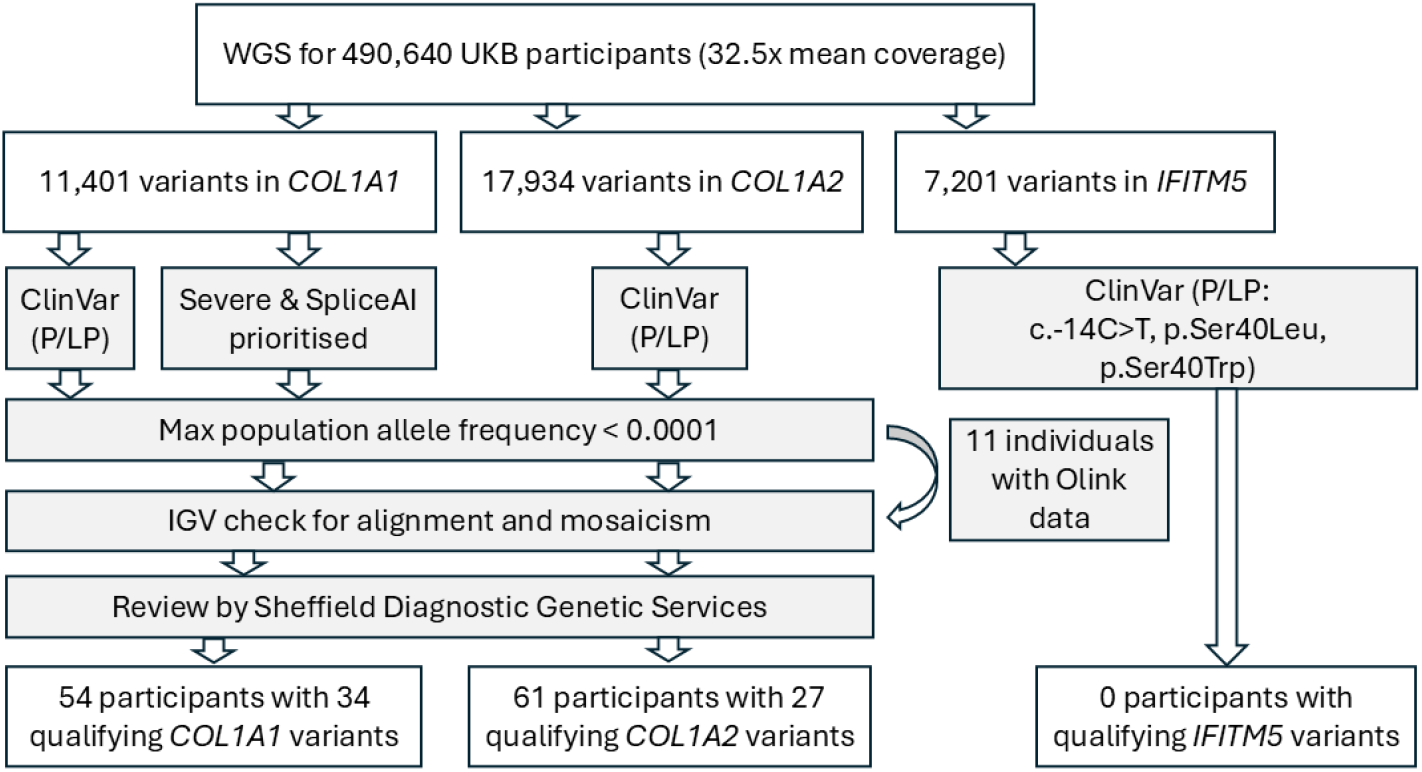
Schematic flow-diagram summarising variant filtering steps in study. Numbers of qualifying variants and numbers of UK Biobank participants harbouring this set of constitutive (likely) pathogenic variants are shown at the bottom. Intermediate filtering steps are highlighted in grey.

- For *COL1A1*, the ClinGen Dosage Sensitivity Curation Working Group has verified that there is sufficient evidence to support haploinsufficiency as a disease mechanism for OI (CCID:006903), with evidence comprising several confirmed pathogenic deletions, frameshifts, start-loss and stop-gains.^20-22^ All pLOF variants were therefore prioritised, regardless of whether the variants had been reported previously in ClinVar (Web resources), or elsewhere. Candidate splice variants were assessed using SpliceAI-visual via a Google Colab notebook.^23^ Previous studies have highlighted the importance of assessing absolute as well as delta SpliceAI scores and exemplar variants in *COL1A1* (c.334-2A>G, c.1056+2T>C, c.1354-12G>A and c.3207+1G>A) demonstrate how a complete view of these scores can assist interpretation (**Figure S1, Note S1**). Specific missense variants in *COL1A1* are also known to cause disease and these were prioritised primarily using ClinVar assertions (pathogenic, P, or likely pathogenic, LP) and with a focus on those that impact the Gly-Xaa-Yaa repeat structure.
- For *COL1A2*, ClinGen has curated it as having no evidence for haploinsufficiency as a disease mechanism (CCID:006904) and over 80% of (L)P variants listed in ClinVar are missense changes, most of which alter Gly-Xaa-Yaa repeats. For this gene, prioritisation was primarily based on ClinVar pathogenicity assessments.
- For *IFITM5*, variant prioritisation was straightforward, as there are only three activating variants known to be pathogenic in this gene. These are a highly recurrent 5′-UTR variant (c.-14C>T, VCV000037143.61) and two missense substitutions involving codon 40: c.119C>T, p.(Ser40Leu) and c.119C>G, p.(Ser40Trp) (VCV000183677.18, VCV000689498.7).

All variants were filtered using a population allele frequency threshold <0.0001 in the gnomAD v3 genomes dataset and a UKB allele count of <10. Genome sequencing reads were visually inspected for all variants using IGV (v2.6.6)^24^ and any lacking sufficient support (e.g. due to poor mapping quality, low read depth, low allelic balance or strand bias) were excluded. Analysis of potential somatic mosaicism involved summing allelic read counts from both exome and genome sequencing datasets. A cumulative binomial test was then performed to evaluate whether the observed allelic fraction differed significantly from the 50% expected for constitutive germline variants. Finally, all qualifying variants were then independently assessed by the Sheffield Diagnostic Genetic Services (SDGS), which currently runs the UK national targeted testing services for OI (Web resources).

### Phenotype analysis

Once the set of high-quality qualifying variants had been finalised, individuals carrying these variants were identified using an in-house custom app available in the research user environment in DNANexus. The R package ukbrapR (Web resources) for working in the UKB Research Analysis Platform was used to identify UKB participants with the ICD-10 code Q78.0 (Osteogenesis Imperfecta). Self-reported illness from the verbal questionnaire data was reviewed for evidence of any bone-related phenotypes. Lastly, for further granularity of phenotype data, we used a custom R script to pull out all available medical records in a strictly chronological order. This included ICD-9, ICD-10, GP records and data from hospital episode statistics.

### Proteomic and Bone Mineral Density assessments

Proteomics data from EDTA blood plasma samples was generated in ∼50,000 individuals in UKB using the Olink technology.^25^ Olink results are available for 2,923 circulating proteins (including COL1A1) as “Normalized Protein eXpression” (NPX) scores in UKB. This measure represents a relative quantification of protein levels, normalised using a log^2^ scale, where a 1-unit difference in NPX equates to a doubling or halving of the protein concentration. NPX scores were downloaded using the Table exporter (DNAnexus) and analysed with RStudio (v4.2). To avoid sample duplicates, the data analysed was from just the first batch (‘instance 0’) only.

Heel Bone Mineral Density (BMD) measurements were also available for 278,754 UKB participants, based on the quantitative ultrasound index through the calcaneus and analysed in a similar way. Standard Polygenic Risk Scores (PRS) for estimated bone mineral density t-score were calculated based entirely from external GWAS data and scores were generated for all UKB participants by Genomics plc, as described.^26^

## Results

### Manual variant curation and exclusion of somatic mosaic variants results in a high-quality set of (likely) pathogenic variants in UKB expected to cause OI types I-IV

Across all UKB participants, the starting number of SNV/indel variants with annotations for the MANE Select transcripts for *COL1A1, COL1A2* and *IFITM5*, was 11,401, 17,934 and 7,201, respectively. Of these 36,536 variants, 74 variants met our initial variant inclusion parameters.

Across both *COL1A1* and *COL1A2*, 16 variants (15 SNVs and one indel) were detected at significantly reduced allelic fractions, highly suggestive of somatic mosaicism (**Table 1**). Ten of these were in *COL1A1* and allelic fractions ranged from 3.4% for c.2299G>A, p.(Gly767Ser) to 35.9% for c.1598G>A, p.(Gly533Asp). For the remaining six variants in *COL1A2*, allelic fractions ranged from 11.6% for c.2314G>A, p.(Gly772Ser) to 32.1% for c.767G>T, p.(Gly256Val). There were four variants where the same variant was seen in both mosaic and non-mosaic states in different participants and so for these, only individuals in whom the variant appeared to be present in a constitutive state were retained (**Tables S1, S2**). Comparison of allelic ratios obtained from exome versus genome sequencing showed a high degree of correlation, with an R^2^ score of 0.60 (**Figure S2**),suggesting these are true mosaic variants; read alignments for representative examples of a mosaic indel (c.2766_2769del) and a mosaic SNV (c.1A>G) in *COL1A1* are shown (**Figure S3A-B)**.

**Table 1:** Summary of (likely) pathogenic variants suspected to be mosaic and removed from penetrance calculations. Alt, number of reads with variant. Ref, number of reads at variant position. VAF, variant allelic fraction. *P*-values were calculated based on the binomial distribution (one-tailed). Protein coordinates are based on ENSP00000225964.6 (*COL1A1*) and ENSP00000297268.6 (*COL1A2*). For each gene, variants are presented in order of decreasing allelic fraction. †, mosaicism is supported by Olink data which showed the COL1A1 protein levels were not reduced (**Figure 3**).

| HGVS (cDNA) | HGVS (protein) | Alt<br>genome | Ref<br>genome | VAF<br>genome | Alt<br>exome | Ref<br>exome | VAF<br>exome | Alt<br>combined | Ref<br>combined | VAF<br>combined | P-value |
| --- | --- | --- | --- | --- | --- | --- | --- | --- | --- | --- | --- |
| *COL1A1* (ENST00000225964.10) |  |  |  |  |  |  |  |  |  |  |  |
| c.1598G>A | p.(Gly533Asp) | 7 | 22 | 31.8% | 30 | 81 | 37.0% | 37 | 103 | 35.9% | 0.0028 |
| c.769G>A | p.(Gly257Arg) | 11 | 28 | 39.3% | 21 | 65 | 32.3% | 32 | 93 | 34.4% | 0.0017 |
| c.1921G>A | p.(Gly641Arg) | 11 | 40 | 27.5% | 17 | 42 | 40.5% | 28 | 82 | 34.1% | 0.0027 |
| c.1A>G† | p.(Met1?) | 12 | 57 | 21.1% | 17 | 48 | 35.4% | 29 | 105 | 27.6% | 3E-06 |
| c.2902G>T | p.(Gly968Ter) | 5 | 27 | 18.5% | 7 | 48 | 14.6% | 12 | 75 | 16.0% | 8E-10 |
| c.904-2A>C | - | 4 | 35 | 11.4% | 10 | 54 | 18.5% | 14 | 89 | 15.7% | 2E-11 |
| c.3082G>A | p.(Gly1028Ser) | 6 | 46 | 13.0% | 10 | 59 | 16.9% | 16 | 105 | 15.2% | 9E-14 |
| c.2766_2769del | p.(Gly923ProfsTer184) | 6 | 36 | 16.7% | 8 | 76 | 10.5% | 14 | 112 | 12.5% | 5E-17 |
| c.369+1G>A | - | 4 | 20 | 20.0% | 2 | 80 | 2.5% | 6 | 100 | 6.0% | 1E-21 |
| c.2299G>A | p.(Gly767Ser) | 3 | 25 | 12.0% | 2 | 122 | 1.6% | 5 | 147 | 3.4% | 3E-36 |
| *COL1A2* (ENST00000297268.11) |  |  |  |  |  |  |  |  |  |  |  |
| c.767G>T | p.(Gly256Val) | 13 | 37 | 35.1% | 14 | 47 | 29.8% | 27 | 84 | 32.1% | 0.0007 |
| c.838G>A | p.(Gly280Ser) | 13 | 28 | 46.4% | 13 | 53 | 24.5% | 26 | 81 | 32.1% | 0.0008 |
| c.70+717A>G | - | 11 | 37 | 29.7% | 0 | 0 | NA | 11 | 37 | 29.7% | 0.0100 |
| c.704G>A | p.(Gly235Asp) | 9 | 34 | 26.5% | 12 | 53 | 22.6% | 21 | 87 | 24.1% | 7E-07 |
| c.767G>A | p.(Gly256Asp) | 10 | 48 | 20.8% | 12 | 49 | 24.5% | 22 | 97 | 22.7% | 3E-08 |
| c.2314G>A | p.(Gly772Ser) | 3 | 25 | 12.0% | 5 | 44 | 11.4% | 8 | 69 | 11.6% | 2E-11 |

Manual review through visual inspection of sequence reads also identified variant co-occurrence, thus avoiding ‘double-counting’ the same participant. Two notable examples of this were c.1602del and c.1604del in *COL1A1*, which were seen *in trans* (**Figure S3C**), whilst c.1A>C and c.2_5del were found *in cis* (**Figure S3D**). In each case, only the better supported variant (c.1602del and c.2_5del) was retained (**Table S1**). Further literature review led to additional exclusions. For instance, pLOF variants in *COL1A2* are linked to an autosomal recessive condition ‘Ehlers-Danlos syndrome, cardiac valvular type’ (MIM: 225320), but not OI. The splice-donor variant NM_000089.4:c.1404+1G>A identified in three UKB participants is a confirmed pathogenic variant (OMIM: 120160.0047) and listed as pathogenic in ClinVar (VCV000017275.5), but only linked to the recessive condition and thus was excluded from our analysis of autosomal dominant OI.

Following the manual curation steps described above, in *COL1A1* there were 34 qualifying variants observed as heterozygous in 54 UKB participants (**Figure 1, Table S1**). Meanwhile in *COL1A2* there were 27 qualifying variants observed as heterozygous in 61 UKB participants (**Figure 1, Table S2**). No (likely) pathogenic CNVs in *COL1A1/2* were identified (**Note S2**). Of the 61 high-quality qualifying variants identified, 47 appeared as singleton alleles in the UKB. In contrast, the most common variants (NM_000089.4:c.3196G>A and NM_000089.4:c.2701G>A) were each seen in nine individuals. There were 29 pLOF or splice variants and 32 missense substitutions, of which 29 altered a Glycine residue in the respective Gly-Xaa-Yaa repeat regions. Of the 61 variants, 45 were SNVs and 16 were indels. CADD scores ranged from 16 (NM_000088.4:c.1354-12G>A) to 49 (NM_000088.4:c.1981C>T; p.Gln661Ter). The majority of qualifying variants (53/61) were listed in ClinVar at the time of last assessment (31 P, 10 LP/P and 10 LP), with two of the retained variants currently listed as ‘conflicting’; for instance NM_000089.4:c.1423G>A (p.Gly475Ser) was LP/P in a previous version (VCV001398959.6), but a single VUS assessment in the current version (VCV001398959.8) moves the overall classification to ‘conflicting’. Given the 3 P/LP classifications and the fact the variant is expected to disrupt the triple helical domain we decided to retain this variant. For *COL1A1*, start-loss is a known disease mechanism^20^ and so an indel involving the start codon (NM_000088.4:c.2_5del) was also retained. Otherwise, variants were widely distributed across both genes (**Table S1 and S2, Figure S4**).

### Despite minimal depletion of OI in UKB, penetrance of pathogenic variants in COL1A1/2 was lower than in clinical cohorts

Extrapolation from the reported prevalence of OI (0.3 - 1.5 per 10,000 individuals)^7,9^ predicts between 15 and 75 individuals with OI amongst ∼500k individuals within UKB. Based on ICD-10 code Q78.0, there were 76 individuals in UKB with OI, a value close to the upper bound of the expected range, consistent with minimal depletion due to “healthy volunteer” selection bias.^27^ Although the most severe forms of childhood-lethal OI would not be expected to be present in an adult cohort, this prevalence suggests that clinical data in UKB is likely to be reasonably complete for the OI phenotype. Nevertheless, for greater certainty we also scrutinised all available electronic health records (including hospital episode statistics and GP records) in a chronological order, searching for the presence of multiple fracture events that could not be explained by environmental factors or other events captured in the records. Individuals with fractures in close temporal proximity to V43.5/V18.4 or W51 codes (driver/cyclist injured in traffic accident, accidental striking against or bumped into by another person) were not considered to be affected.

Using all relevant health records to assess OI phenotypes, the estimated penetrance across UKB for adult carriers of (likely) pathogenic variants in *COL1A1* was 40.7% (22/54), with most of these positive cases from ICD-10 coding (**Figure 2A**). Restricting the set of variants to include just pLOF variants in *COL1A1*, penetrance estimates increased to 73.1% (19/26), whilst the penetrance seen for missense *COL1A1* variants dropped to 10.7% (3/28) (**Figure 2B, C**). In contrast, estimated penetrance for carriers of (likely) pathogenic variants in *COL1A2* was only 21.3% (13/61), with just five coming from ICD-10 codes (**Figure 2D**). These results are substantially decreased from penetrance estimates based on clinical cohorts, where penetrance for heterozygous *COL1A1* or *COL1A2* pathogenic variants has been estimated at close to 100%.^8^ Most ICD-10 diagnoses in carrier individuals occurred between the ages of 40 and 60 (**Figure 2E**).

**Figure 2:**
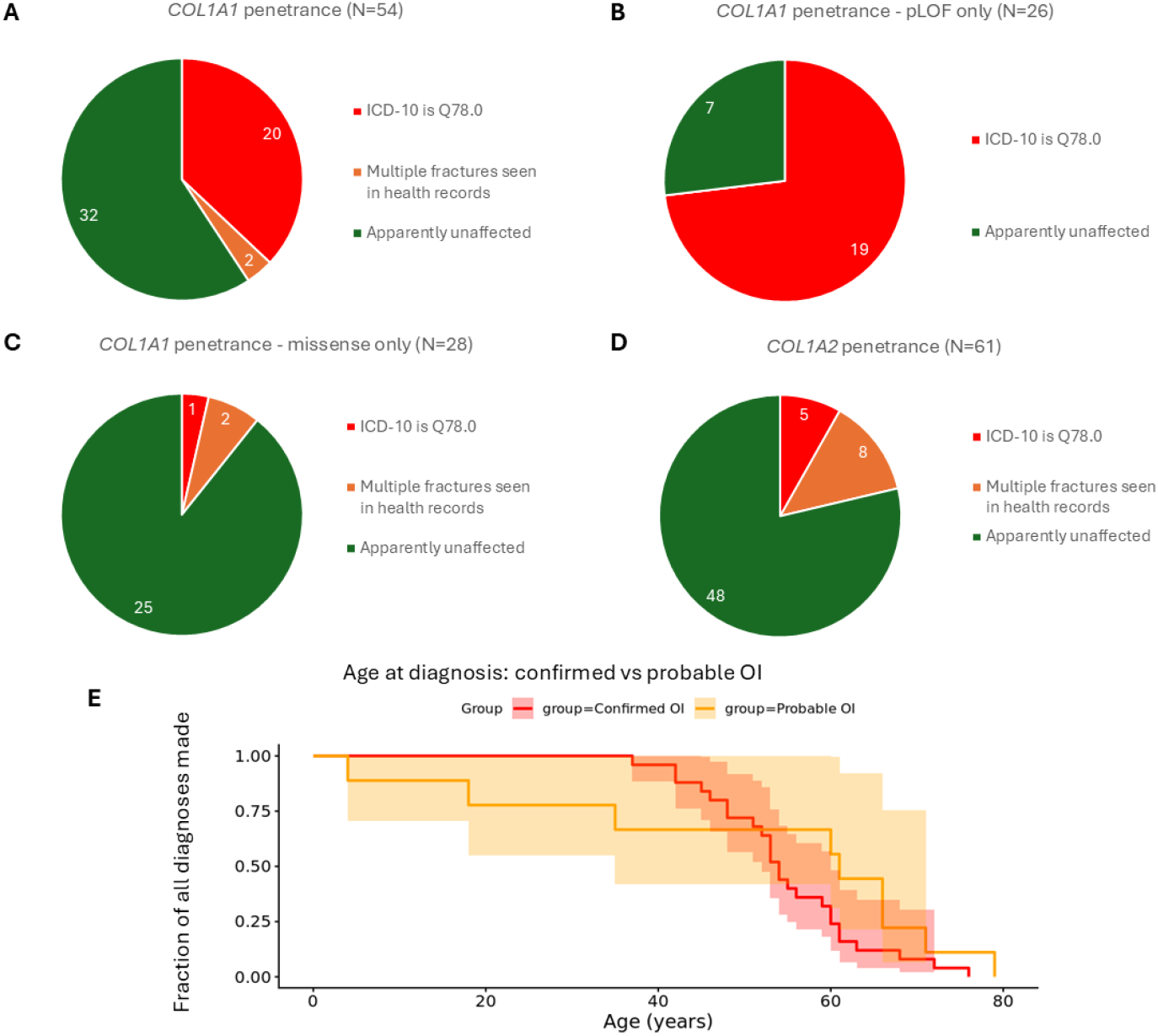
Incomplete penetrance and age at diagnosis for *COL1A1* and *COL1A2* variant carriers. A) Penetrance results for *COL1A1*. The red segment in the pie chart represents individuals with a qualifying variant and where ICD-10 codes include Q78.0. Orange segments denote variant carriers where there is not Q78.0 but where the individual has had multiple fractures. Green segments denote individuals with a variant but where that individual has neither the Q78.0 code nor evidence of multiple fractures. Penetrance estimates were then recalculated using *COL1A1* variants subdivided into B) those that are pLOF (including splice variants) and C) those which are missense. D) Penetrance results for carriers of qualifying *COL1A2* variants. E) Plot showing the approximate ages at which the ICD-10 code Q78.0 was assigned for the 25 variant carriers (range 37-76, shown in red), compared to the 9 individuals where affected status was inferred, based on presence of multiple fractures (4-79 years, orange). For one individual, the date of second fracture was not available and this individual was omitted from the plot. Shaded bands represent 95% confidence intervals.

### Proteomic data aids variant curation and supports functional effect of pLOF variants in COL1A1

At the time of analysis, there were 52,155 UKB participants with Olink circulating protein data for COL1A1. The 25th centile for NPX score was at -0.1847 whilst the 1st centile was at -0.645, with just 522/52,155 results lying below this second threshold. Of the participants with potentially qualifying variants in *COL1A1*, 11 had Olink data available and, by chance, these all involved pLOF variants. Reassuringly, the *COL1A1* NPX scores for this set of individuals mostly clustered around -1, consistent with heterozygous pLOF variants resulting in a 50% drop in protein levels (**Figure 3**). Interestingly, a high NPX score for a known pathogenic start-loss variant in one of these individuals helped support its exclusion due to likely somatic mosaicism (**Table 1, Figure S3B**). Of the remaining 10 individuals with pLOF variants in *COL1A1* and Olink results, the levels of α1 collagen protein were below the 1^st^ centile in nine (90%), of whom eight also had the ICD-10 code for OI (Q78.0).

**Figure 3:**
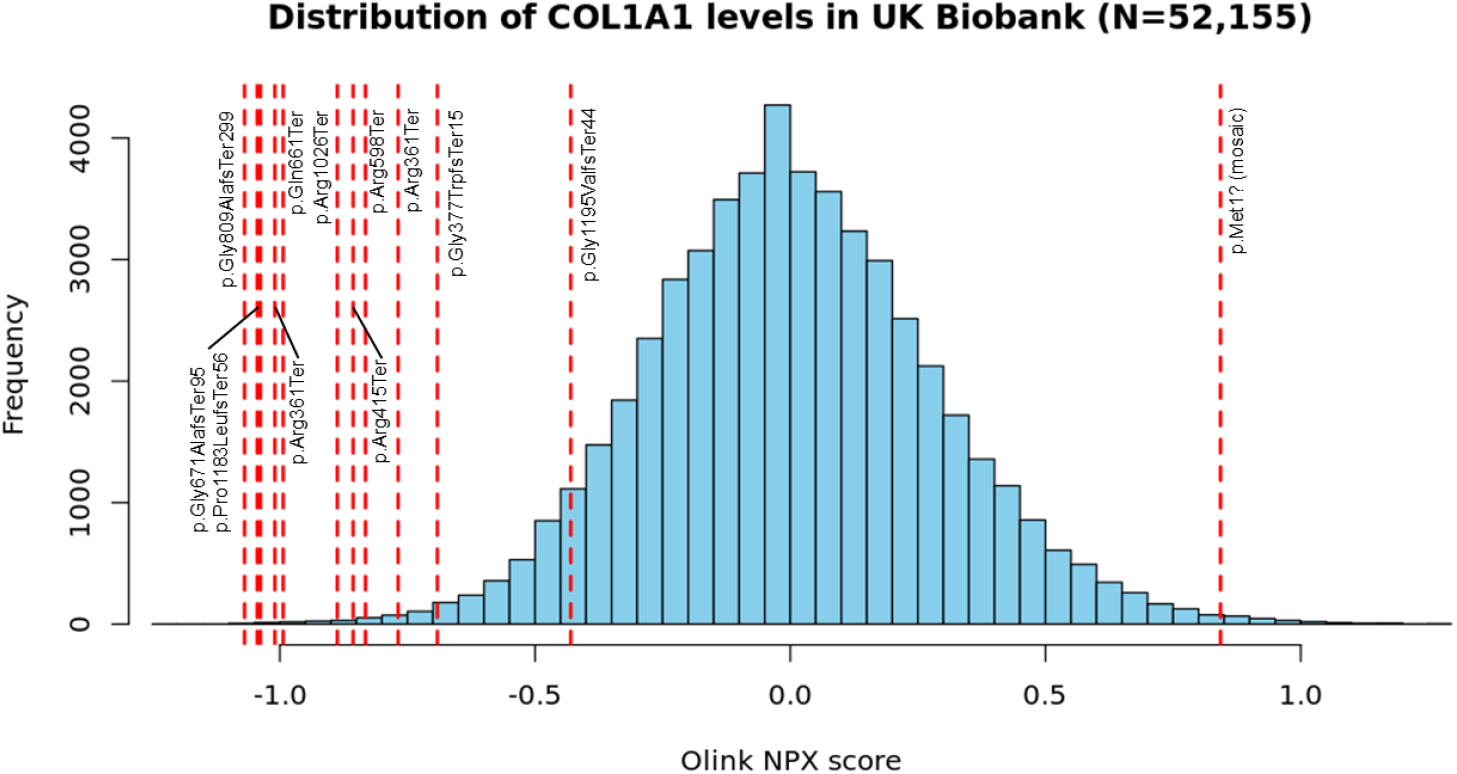
Proteomic data for *COL1A1*/*COL1A2* variant carriers. Olink blood proteomics data from UK Biobank supports a deleterious effect of pLOF variants on protein levels. Histogram plot created with RStudio (v4.4) and data for individuals with heterozygous *COL1A1* pLOF variants are labelled. The c.1081C>T, p.(Arg361Ter) variant was detected in two UB biobank participants with Olink data availability. The p.Pro1183LeufsTer56 variant was detected in an individual for whom only exome data was available and so was not included in our penetrance calculations. The c.1A>G (p.Met1?) variant was only present in 27.6% of reads (**Figure S3B, Table 1**) and so is likely mosaic, explaining the lack of any decrease in relative protein levels. The p.Gly1195ValfsTer44 variant is the most terminal of all variants shown and thus the protein may retain some stability – this could explain why there is only a partial reduction of protein levels for that sample. All 52,155 datasets are from instance 0 and the mean Normalized Protein eXpression (NPX) value was 0.0038.

A reciprocal analysis was performed starting with all 76 individuals in UKB with ICD-10-defined OI. Of these, only 13 had Olink data available and of those, nine had an NPX score below the 1^st^ centile. From these nine individuals, eight harboured a qualifying *COL1A1* variant detected by genome sequencing (i.e. the same positive variants/individuals mentioned above). The only additional individual in UKB with Q78.0 and a reduced level of α1 collagen but without a *COL1A1* variant in the genome sequencing data was subsequently found to carry a c.3548del, p.(Pro1183LeufsTer56) variant detected via the exome sequencing pipeline (genome sequencing data unavailable for this participant). These results highlight that the combination of a clinical OI diagnosis and an Olink-confirmed reduction of α1 collagen is highly specific for the presence of a pathogenic *COL1A1* pLOF variant.

### Correlation with bone mineral density supports low penetrance of missense variants in COL1A2

With the exception of variants that disrupt the type I procollagen C-propeptide cleavage site which result in raised BMD^28,29^, OI-associated variants in *COL1A1* and *COL1A2* are typically associated with decreased BMD and the degree of change depends on whether the variant confers a qualitative or quantitative change to protein levels. Although BMD measures of clinical relevance to OI are best taken in the lumbar spine, such data are only available for 45,522 UKB participants. We therefore used the normalised heel BMD data which was available for over half of participants (N=278,975, mean of -0.337). For individuals who had both measurements (**Figure S5A**), heel BMD showed a highly significant correlation with L1-L4 BMD scores (*P* < 2.2×10^-16^), but with an R^2^ of only 0.138 (**Figure S5B**). For the 27/54 (50%) individuals with qualifying *COL1A1* variants where heel BMD was available, the mean BMD was significantly reduced (-1.315; *P*=7.22×10^-5^). The mean heel BMD was also reduced in 38/61 (62%) individuals with *COL1A2* variants where BMD was available (-0.846; *P*=0.016), although to a lesser extent.

Variant carriers were then split into clinically affected and unaffected subcategories. Comparisons of BMD measurements between these groups (**Figure 4**) revealed no significant difference between BMD in clinically unaffected individuals with (likely) pathogenic *COL1A2* variants and UKB participants without (likely) pathogenic *COL1A2* variants (*P*=0.808). In contrast, for affected individuals with (likely) pathogenic *COL1A2* variants, mean BMD values decreased from -0.34 to -1.99 (*P*<0.001). For *COL1A1*, the mean BMD for clinically unaffected carriers (-1.23) was intermediate between non-carriers (-0.34) and affected carriers (-1.81). These changes in BMD in clinically affected individuals due to a single rare variant were over twice those seen when comparing mean BMD results in UKB participants stratified by PRS, where a 0.68 difference was seen comparing quantile 1 to quantile 5 (**Figure S6**).

**Figure 4:**
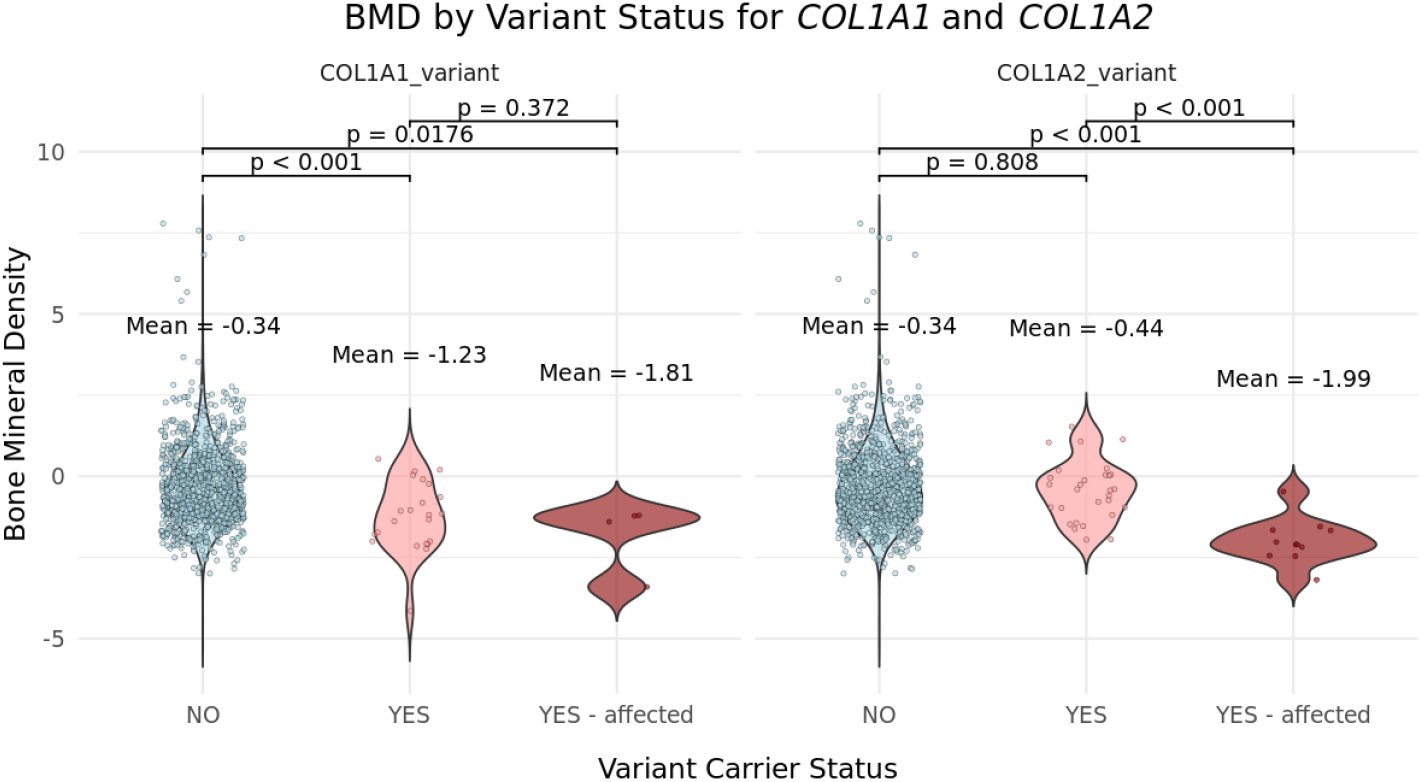
Bone mineral density (BMD) data for *COL1A1*/*COL1A2* variant carriers. Violin plots show the distribution of heel BMD values across 278,975 individuals from the UK Biobank. BMD results for non-carriers (‘NO’, shown in blue) are compared to individuals with qualifying *COL1A1* variants (N=27) and *COL1A2* variants (N=38). Variant carriers are split into those without a clinical phenotype (‘Yes’, pink) and those with a clinical phenotype (‘Yes – affected’, red). For both clinically affected groups, the BMD is significantly lower than across the UK Biobank as a whole. For *COL1A1*, BMD was also significantly reduced for variants carriers who did not have a clinical phenotype. Statistical comparisons between groups were performed using the Wilcoxon rank-sum test in RStudio. To aid visualisation, BMD results for non-carriers were randomly down-sampled before data points were plotted with the jitter function.

One possible hypothesis for the incomplete penetrance is that increased PRS scores for BMD could result in a milder (i.e. adult-onset) or subclinical presentations for carriers of rare pathogenic rare variants. However, there was no significance difference (*P*>0.1) in the BMD PRS between the *COL1A1/2* variant carriers and the rest of UKB (**Figure S7**).

### No pathogenic variants were identified in IFITM5, supporting extreme rarity and high penetrance of OI type V

None of the three (likely) pathogenic variants in *IFITM5* were identified in UKB. The lack of suspected pathogenic genotypes for this gene across such a large cohort is consistent with the extreme rarity of OI type V and/or a suspected high penetrance combined with the typically acute and specific phenotypic presentation. Although another missense variant at the same amino acid position, c.118T>C p.(Ser40Pro), was identified in two heterozygous individuals, this is a VUS in ClinVar (VCV003626519.1). Based on the ACMG/AMP criteria,^30^ PM5 (‘novel missense at the same position as a pathogenic variant’) should only be applied with caution, especially if a gain-of-function mechanism is suspected. Of note, the two (likely) pathogenic variants involve amino-acid substitutions considered to be more deleterious (based on Grantham score) and also predict the same new splice-donor site within exon 1, 3 bp downstream (DG=0.64/0.14), which is not the case for c.118T>C. Given the lack of evidence supporting pathogenicity, this variant was not evaluated further.

## Discussion

Rare variants in *COL1A1* and *COL1A2* that cause bone fragility are typically detected in around 80-90% of individuals with OI.^7,9,12^ However, there is ongoing debate about whether these genes should be included in genomic NBS programs as globally they are tested in under half of such projects. Of the panels assessed by Minten *et al, COL1A1* was present in 12/27 and *COL1A2* in 13/27.^5^ Furthermore, modelling of the NBS variant prioritisation strategy from the UK’s Generation Study identified *COL1A1* and *COL1A2* as outliers in terms of the large numbers of variants prioritised,^31^ and so care needs to be taken to limit reporting of variants incorrectly associated with disease, or those with low penetrance. Here, we used genome sequence data from half a million adults from UKB and show that, for a curated set of (likely) pathogenic variants in *COL1A1* and *COL1A2*, there is incomplete penetrance (∼40% and ∼20%, respectively) with respect to a clinical diagnosis of OI and/or the presence of multiple fractures in the available electronic health records. This is in contrast to the literature, as although variable severity is widely reported, penetrance has previously been considered to be near complete.^32^ Our result therefore mirror other studies which suggest that disease penetrance estimates in the literature can often be biased as ascertainment has historically used clinical, rather than population cohorts.^3^

The OI phenotype resulting from missense variants in *COL1A1* and *COL1A2* in particular is typically thought to be more severe than OI due to *COL1A1* haploinsufficiency.^7,32^ This is due to the dominant-negative effect certain missense changes can have on the triple helix structure of collagen. Our penetrance estimates were therefore initially counterintuitive, as the penetrance for pLOF and splicing variants in *COL1A1* was higher (73.1%) than missense changes in the same gene (10.7%) or in *COL1A2* (21.3%). There are several possible explanations for this finding. Firstly, it is much more reliable to infer that frameshift and stop-gain variants result in loss-of-function than it is to predict the functional consequence of a missense alteration, and therefore a proportion of the included missense variants may not be truly pathogenic. Secondly, lower than anticipated penetrance may be due to the absence of severe childhood-onset OI phenotypes in UKB, due to healthy volunteer bias.^27^ Taken together, these explanations would amplify the effect of any remaining missense variants in UKB which may have been retained due to incorrect assertions in ClinVar. We also suspect that a lack of suitably matched controls in OI studies and a historical over-reliance on the Gly-Xaa-Yaa rule for variant interpretation has led to a self-propagating issue of overreporting glycine-altering candidate variants. To combat the potential risks of falsely reporting missense variants detected by genomic NBS, there will need to be meticulous curation that assesses the latest up-to-date literature and integrates clinical information with data from large population cohorts, complemented by high-throughput functional testing.^33^ In this study, we sought to mitigate the possibility of inaccurate variant curation through additional scrutinization of each variant by the national OI testing laboratory in Sheffield. Examples of poor or outdated curation in ClinVar are not uncommon (**Note S3**) and assertions based on single submissions should be treated with caution. However, widespread efforts by ClinGen curation teams to flag suspicious or conflicting assertions in ClinVar are ongoing.^34,35^

In contrast to the two α-collagen genes, *IFITM5* was only included in 5/27 of the genomic NBS panels assessed by Minten *et al*.^5^ In this study, no pathogenic variants in *IFITM5 were* identified in UKB. This lack of pathogenic genotypes across such a large cohort is consistent with the rarity of OI type V and/or a suspected high penetrance combined with the typically acute childhood phenotypic presentation. Survivor bias likely accounts for the absence of individuals with such variants in this adult cohort. In contrast, we note that five individuals with *de novo* pathogenic NM_001025295.3:c.-14C>T variant were detected in the 100kGP,^36^ using a similar methodology. Given this relative absence of pathogenic variants in UKB, our results suggest that inclusion of this gene in genomics NBS would likely achieve a high specificity.

Proteomic data is currently available for around 10% of UKB participants and COL1A1 is one of the 2,923 proteins tested on the Olink platform. This enabled us to confirm a detrimental effect on protein levels for the group of pLOF variants (**Figure 3**). This data also supported the exclusion of a mosaic start-gain variant that was seen in 27.6% of reads (**Figure S3B**). Unfortunately, Olink data was unavailable for missense variant carriers as such data would be especially valuable for identifying missense variants that impact protein stability.

Nevertheless, these results highlight the value of complementary proteomic studies for enhancing penetrance studies. Future studies should investigate the wider value of generating companion proteomic data to supplement genomic NBS.

BMD measurements for the lumbar spine are typically the most relevant for assessment of OI. However, this data was only available for <10% individuals, whereas heel BMD was available for >50% of all UKB participants and correlated well with lumbar BMD. We found that the group of individuals with qualifying *COL1A1* variants but no relevant ICD-10 code or evidence for multiple fractures also had a significant BMD decrease. This suggests that a variant may be ‘non-penetrant’ in terms of clinical phenotype but may still have a subclinical effect on bone density and highlights that estimates of penetrance are highly dependent on the outcome measure being assessed. In contrast, BMD data supported low penetrance for *COL1A2* variants as variant carriers with no relevant clinical findings had similar BMDs to the non-carrier group (**Figure 4**).

Several case reports describe (likely) pathogenic *COL1A1/2* variants in individuals with somatic mosaicism and in some cases these findings help explain intra-familial differences in OI expressivity.^37-39^ Where mosaicism results in a milder skeletal phenotype, it is thought that variants still need to be present in >75% of osteoblasts.^38^ In contrast, where mosaicism is confined to blood, this would have little impact on bone development and such variants may potentially contribute towards incorrect assertions of low penetrance. In a population context where testing is limited to a single blood sample, it is impossible to assess relative tissue distributions. It is therefore prudent for such variants that appear mosaic in blood to be removed before performing any penetrance calculations. In a previous study, we suggested an allelic balance of 27% as an appropriate cutoff which achieves a suitable balance between sensitivity and specificity for detecting somatic mosaicism.^40^ In penetrance studies, specificity is arguably more important than sensitivity and given that genome sequencing in UKB was at a mean coverage of only 32.5x, detection of somatic mosaicism would be limited. However, by combining this data with the higher coverage exome sequencing, we were able to increase specificity and remove several variants with allelic fractions of 27-40% (**Table 1**). Examples of retained variants not quite passing the significance threshold for mosaicism are presented in **Note S4**. Combining likely mosaic with false-positive variant calls, a total of 24 *COL1A1* variants were excluded. Without this manual review and systematic mosaicism analysis, this ∼70% increase in qualifying variants would have a major impact on the accuracy of downstream penetrance estimates. Whilst some of these variants could have been filtered using automated QC filtering approaches, this could impact sensitivity and so these results highlight the importance of undertaking a careful manual curation of variants.^41^ Manual review also identified examples of complex multi-nucleotide variants which were called as separate indel/SNVs (**Figure S3D**); care must be taken to avoid ‘double-counting’ for this class of variant.

The present study highlights several generalizable lessons that supplement our previous guidance about estimation of disease penetrance.^41^ For instance, four cases with qualifying variants in *COL1A1* (p.Gly1079Ser, p.Arg312Cys, p.Arg1014Cys and p.Pro535LeufsTer6) had multiple fractures reported and initially these were considered to be likely affected individuals. However, closer scrutiny revealed that all fractures occurred within a short time frame and were concomitant with road traffic collisions or other physical accidents. This suggested the fractures may not be due to a genetic bone fragility condition and as no other evidence for bone fragility was identified, the coding for these individuals was switched to unaffected. These cases illustrate how systematic, chronological presentation of complete electronic clinical records can enhance the study of rare disease.

Although the UKB research analysis platform provides a wealth of clinical records to researchers, it is hard to estimate the effects of missing or incomplete clinical data. This is a major limitation of population cohort studies, as compared to clinically ascertained rare-disease longitudinal cohort studies, which are themselves limited by ascertainment bias. However, given the relatively high penetrance for pLOF variants in *COL1A1* (>70%), this is unlikely to be a major factor. A recent study that studied 91 diseases linked to haploinsufficiency shows surprising levels of incomplete penetrance for pLOF variants in haploinsufficiency disease genes and suggests this may be driven by residual allelic activity, rather than by missing data.^4^ Moreover, in the context of NBS, where potential disease-causing variants would be reported within the first month of life, the presence of known pathogenic variants in adults >40 years of age who were able to volunteer for a population study gives pause for thought.

This finding implies that either the variants are incompletely penetrant and could result in substantial overdiagnosis if reported in the absence of a phenotype, or that individuals have milder or sub-clinical forms of disease that may be unnecessary to detect in newborns.

In conclusion, our study using genome sequencing results from a clinically-unselected UK population cohort of ∼500,000 adults highlights the importance of using population data to inform population screening efforts and the value of integrating proteomic/biometric data alongside genomic data. Our results suggest the penetrance of OI in the general population is lower than for clinically ascertained cohorts, which is only explained in part by depletion of severe early-onset forms of the disease. Furthermore, particularly for missense variants that alter the Gly-Xaa-Yaa repeat structure in the α-collagen genes, the low penetrance suggests that over-reliance on current ClinVar assertions to support pathogenicity may also contribute to the danger of overstating OI risk in genomic NBS screening efforts.

## Data Availability

Data from UK Biobank cannot be shared publicly because of data availability and data return policies. Data are available from the UK Biobank for researchers who meet the criteria for access to datasets to UK Biobank

https://www.ukbiobank.ac.uk

## Web resources

ClinVar, www.ncbi.nlm.nih.gov/clinvar

NHS Genomic Medicine Service Test Directory, www.england.nhs.uk/publication/national-genomic-test-directories

PanelApp, https://panelapp.genomicsengland.co.uk

PanelApp Australia, https://panelapp-aus.org

Sheffield Diagnostic Genetic Services, www.sheffieldchildrens.nhs.uk/services/metabolic-bone-disease-service

ukbrapR, an R package for working in the UK Biobank Research Analysis Platform, https://lcpilling.github.io/ukbrapR

VEP Annotation Process using Docker, https://github.com/drarwood/vep_ukb_aou_docker

## Data and code availability

Data from UK Biobank cannot be shared publicly because of data availability and data return policies. Data are available from the UK Biobank for researchers who meet the criteria for access to datasets to UK Biobank (www.ukbiobank.ac.uk). Code used as part of this study is either listed in the Web resources, or is to be described elsewhere. Further details are available on request.

## Acknowledgments

This research has been conducted using the UK Biobank Resource under Application Number 103356 and uses data provided by patients and collected by the NHS as part of their care and support. The authors would like to acknowledge the use of the University of Exeter High-Performance Computing (HPC) facility in carrying out this work. This work was supported by the Medical Research Council [MR/X021351/1] and the National Institute for Health and Care Research Exeter Biomedical Research Centre. The views expressed are those of the author(s) and not necessarily those of the NIHR or the Department of Health and Social Care.

## Author Contribution Statement

A.T.P analysed data and wrote the manuscript. E.L.B., L.J. and C.F.W. conceived and supervised the study.

R.N.B. provided statistical advice and assisted with variant analysis. D.B. and M.B helped with variant curation, clinical phenotypes in OI and contributed to manuscript writing. J.F. and T.H. wrote custom scripts for data analysis.

## Ethical Approval

UK Biobank protocols were approved by the National Research Ethics Service Committee.

## Declaration of generative AI and AI-assisted technologies in the manuscript preparation process

During the preparation of this work, the authors used ChatGPT to assist with grammar correction and improving readability. The authors reviewed and revised all content generated by the tool and take full responsibility for the final manuscript.

