## Supplemental for "Reduced penetrance of *COL1A1/2* pathogenic variants linked with osteogenesis imperfecta: analysis of a large population cohort"

### Supplementary Material

**Note S1: Exemplar variants highlighting difficulties of assessing splice variants**

Given the high proportion of non-terminal exons that are in-frame in *COL1A1* (46/49) and *COL1A2* (47/50), the functional effects of splice variants are hard to predict and so a second limitation of our study was that no RNA studies were performed. We therefore relied on SpliceAI predictions (absolute scores generated using SpliceAI-visual[1]), in combination with population allele frequencies and evidence from database/literature, and a total of six splice variants were retained (**Tables S1, S2**). A notable example is the c.334-2A>G splice variant in *COL1A1* which disrupts the acceptor site for exon 4, but also predicts the introduction of a new acceptor site, shortening the exon by 9bp (**Figure S1**). As well as being in-frame, this would retain the Gly-Xaa-Yaa pattern and hence the functional consequence of this alteration is uncertain. Together with its absence from ClinVar, this meant that c.334-2A>G was discarded. In contrast, a nearby c.334-9A>G (four P/LP submissions in ClinVar; VCV000569955.17), lies outside the splice region prioritised by many *in silico* pipelines but predicts an 8bp extension to the same exon and is highly likely to result in loss of function and lead to OI type I.

**Note S2: Absence of likely pathogenic CNVs in *COL1A1/COL1A2* across UK Biobank**

A custom R script was also used to prioritise CNVs that overlap any exonic sequence of *COL1A1* and *COL1A2*. To help reduce false positive calls, only CNVs with a minimum size of >10 kb were retained. Viewing read alignments using IGV v2.6.6 was done in order to verity CNV calls and confirm precise structures, as it is known that approaching 20% of rare duplications involve atypical structures.[2]

In *COL1A1,* we identified 5 individuals with 4 different duplications of between 32.6 kb and 16.5 Mb in size. Of these, 3 were full gene duplications. The 32.6 kb duplication, present in two individuals, involved all but the first exon of *COL1A1*. Review of breakpoints indicated that none of these CNVs were likely to be deleterious to gene function and triplosensitivity is not a known disease mechanism for this gene, so these duplications were not prioritised. No exonic deletions were identified. For *COL1A2*, CNVs ranged in size from 202 kb to 2.1 Mb and included two whole gene duplications, a whole gene deletion and another deletion involving exons 1-6. Most reported pathogenic *COL1A2* mutations result in a structurally abnormal protein product and pathogenesis is via a dominant-negative mechanism.[3] CNVs therefore need to be evaluated carefully and only those that can result in a structurally abnormal protein should be prioritised. As deletions were all likely to result in total loss of protein, these were not included for further analysis in our penetrance studies. CNVs involving *IFITM5* are not expected to cause OI and so were not evaluated.

A limitation of this study is that although we assessed CNVs down to 10kb in size, copy neutral SVs were not assessed. However, studies have shown that inversions can potentially play a role in only around 1/700 families with a rare genetic condition[4] and so non-detection of this variant class is unlikely to have any large impact on overall penetrance estimates.

**Note S3: ClinVar annotations should be treated with extreme caution**

During the course of this work, we identified several cases where ClinVar assessments were incorrect or misleading. Exemplars of this include a NM_000089.4(COL1A2):c.1009G>A (p.Gly337Ser) (accession VCV000425643.37 where 9/10 assessments are Pathogenic (P) but the final one is VUS and so the top-level assessment is downgraded to conflicting. This has now been fixed in the latest version of this variant record (VCV000425643.52). In general, ClinVar users should also be wary of single LP assessments that were submitted before the availability of large population aggregate data such as gnomAD and ExAC – for example, there is a single LP assessment for NM_000088.4(COL1A1):c.2594G>A (p.Arg865His) from 2011 but the allele count (AC) in gnomAD v4.1.0 is 30 and the variant does not disrupt a glycine residue (VCV000035914.7). Again, the updated record for this variant shows additional assessments have been more cautious and include 2 VUS and one benign assertion (VCV000035914.14). Another example of a poorly curated ClinVar entry, first uploaded to ClinVar in October 2020, is NM_000088.4(COL1A1):c.992C>T (p.Ala331Val) (VCV000870110.33). This was coded as LP (SCV001244826.2) but the submitter comments that the variant is co-transmitted (i.e. *in cis*) with a much more deleterious allele (c.3008del) and so the LP assessment is likely linked to the haplotype, not the specific variant being assessed. Lying *in cis* with a known pathogenic variant is evidence against pathogenicity in the ACMG analysis framework (BP2).[5] Although this record was updated April 2025, this assertion has yet to be downgraded.

These examples highlight that ClinVar assessments do change over time and so ideally the latest versions should always be used. Assessments can even change for the same submitter for same variant. As an example of this, we highlight an LP variant that is assessed by GeneDx as LP (VCV000420060.6) but then downgraded to a VUS in the next version (VCV000420060.7). Whist an initial study showed co-segregation with atypical OI for NM_000088.4(COL1A1):c.3196C>T (p.Arg1066Cys) in combination with a weak functional effect,[6] there was less delay in helix formation than would be expected for a Gly substitution. A later study found the variant was inherited from a clinically unaffected father in three siblings with OI who also had a maternally-inherited pathogenic variant *in trans* – the authors hypothesized that it may contribute to the children's phenotype.[7] However, now with the access to the latest population databases (e.g. the allele count for this variant in gnomAD v4.1.0 is 71), in hindsight it is clear that this variant is unlikely to represent a fully penetrant disease associated variant and the conclusions of these earlier studies were speculative. These examples highlight that the ClinVar database is in constant flux and although constantly improving, it likely still contains many inaccurate or questionable assertions.

**Note S4: Additional examples of variants with a suspicion of mosaicism**

Despite significant efforts to exclude mosaic variants, a limitation of our study remains that genetic mosaicism where the true allelic fraction was 40-50% in blood would have remained undetectable. For instance, using the combined data, c.3235G>A (p.Gly1079Ser) in *COL1A1* (pathogenic in ClinVar, VCV000017322.14) was detected in 45/104 reads, whilst c.1576G>A (p.Gly526Arg) in *COL1A2* (pathogenic/likely pathogenic in ClinVar, VCV000521008.33) was detected in 32/79 reads. These both just missed the cutoff for statistical significance (43.3%, *P*-val=0.1011; 40.5%, *P*-val=0.0573, respectively) and so were retained. At higher sequencing depth, it would likely be possible to determine whether these reduced allelic fractions represent genuine somatic mosaicism or simply stochastic deviations from 50%. Inheritance studies to confirm or refute genetic mosaicism are not feasible in large biobank-scale studies. Mosaicism in the context of NBS would likely be less of an issue as levels of somatic mosaicism increase during an individual's lifetime.[8] Unlike genes linked to hematopoiesis such as *DNMT3A* and *TET2*,[9] the genes assessed here (*COL1A1, COL1A2* and *IFITM5*) are not known to be hotspots for somatic mosaicism.

**Table S1:** Qualifying variants in *COL1A1* and number of carriers. *Provided as separate xlsx file.*

**Table S2:** Qualifying variants in *COL1A2* and number of carriers. *Provided as separate xlsx file.*

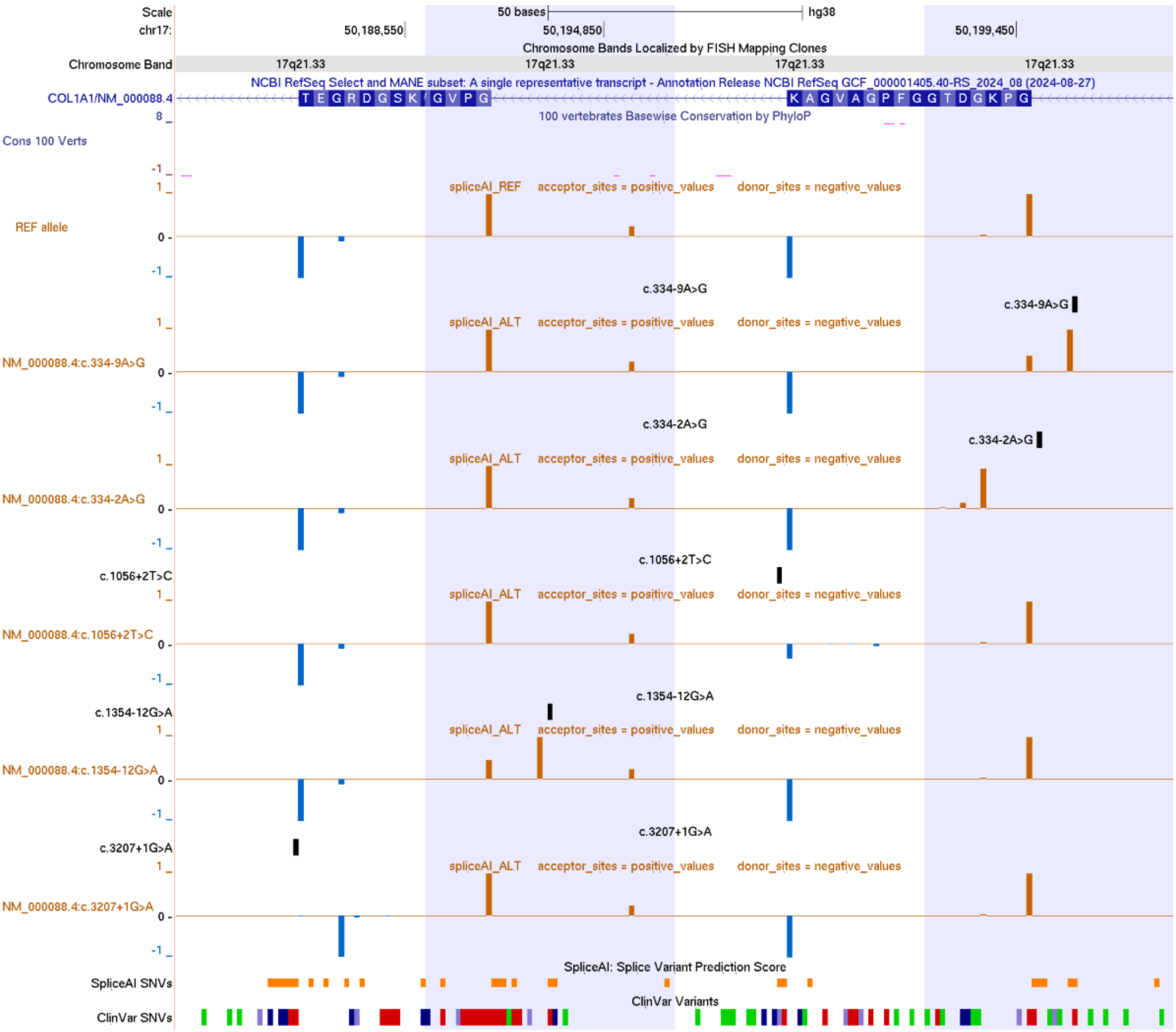


**Figure S1:** SpliceAI-visual plots can assist variant interpretation. Multi-region view on GRCh38 involving four 50 bp windows for exon 43 (chr17:50188505-50188554), exon 21 (chr17:50194815-50194864), exon 16 (chr17:50195900-50195949) and exon 4 (chr17:50199432-50199481). UCSC browser graphic shows absolute SpliceAI scores generated using SpliceAI-visual (https://tinyurl.com/spliceai-visual) and loaded to UCSC as BedGraph files. The c.334-2A>G variant introduces a new in-frame acceptor and shortens exon 4 by 9bp which retains Gly-Xaa-Yaa pattern. In contrast, the nearby c.334-9A>G which is known to be pathogenic (VCV000569955.17, not seen in the present study) results in an 8 bp extension of exon 4. The c.1056+2T>C variant has an allele count in UKB of 19 and likely results in leaky in-frame exon skipping which retains the Gly-Xaa-Yaa pattern. The c.1354-12G>A variant introduces a new acceptor site and is predicted to result in a 10 bp extension of exon 21, causing a frameshift. The c.3207+1G>A variant prediction is that exon 43 shortened by 8bp so leads to frameshift. Based on the above information, c.334-2A>G and c.1056+2T>C were discarded whilst c.1354-12G>A and c.3207+1G>A were retained. An interactive UCSC session is available at https://genome.ucsc.edu/s/ExeterGenetics/COL1A1_splice_figure.

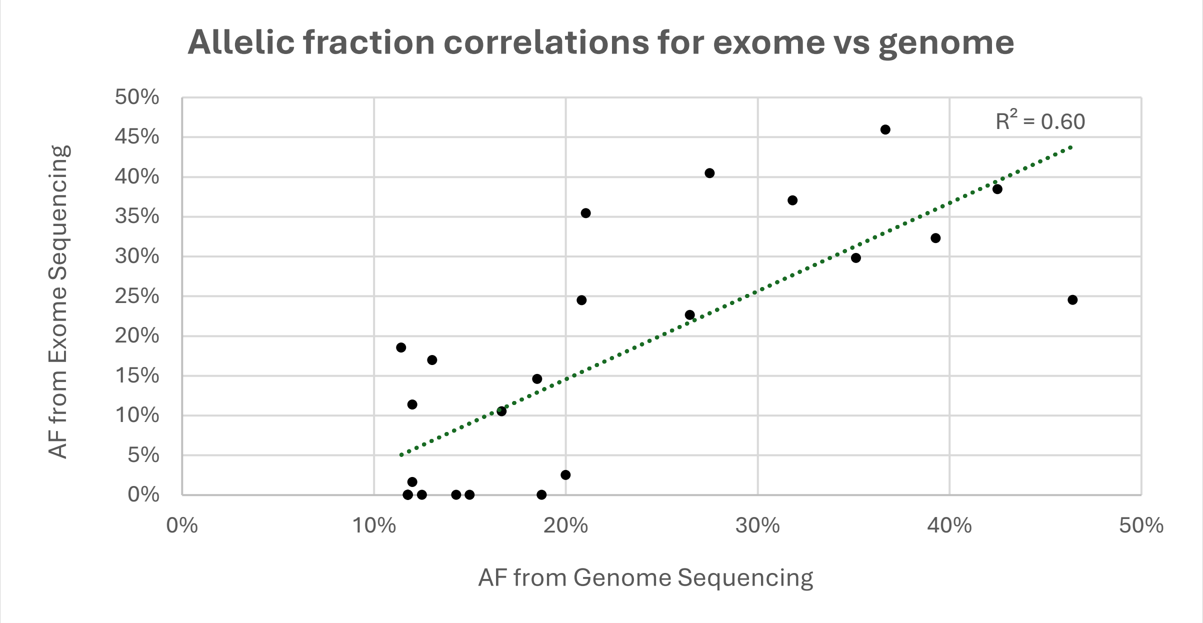


**Figure S2:** Correlation of allelic fractions between exome and genome sequencing data. c.70+717A>G in *COL1A2* was not included in the plot as exome data was uninformative. The other 15 variants from **Table 1** are included, alongside 2 variants which just missed the binomial cutoff (*P*-val > 0.05, **Note S4**) and 8 variants with zero supporting reads in exomes (4 data points overlap, all with 11.8% reads in genome sequencing), suggesting that these variants were false-positive calls in the genome sequencing data.

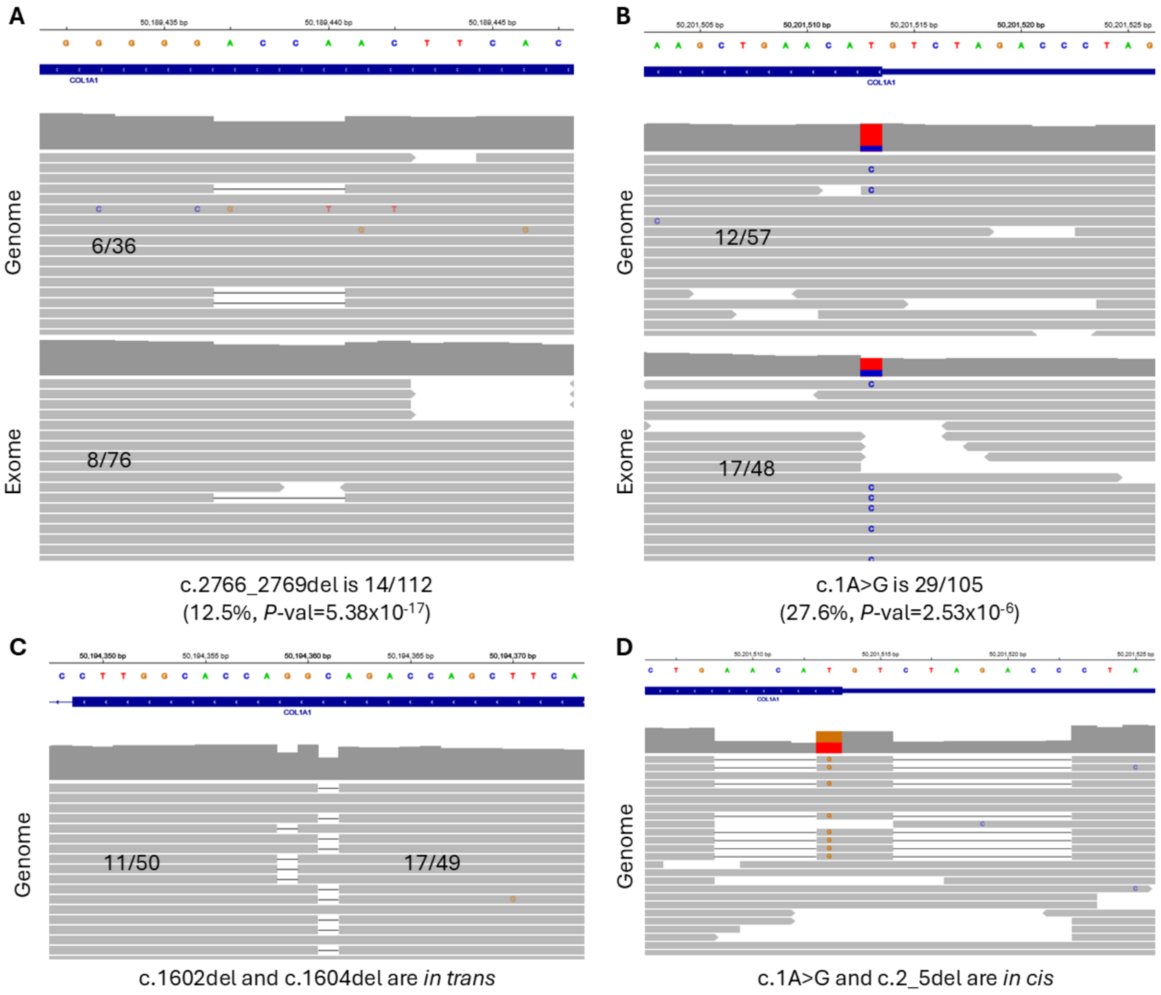


**Figure S3:** Examples of variants excluded following manual variant curation. A) The c.2766_2769del variant appears mosaic based on the combined read count distribution from exome and genome sequencing. B) The start-loss c.1A>G variant also appears mosaic given the combined read count data. C). Although c.1602del and c.1604del appear to lie *in trans*, the presence of some wild-type reads suggests that at least one of these deletions is likely mosaic. No informative SNVs were close enough to allow haplotype studies. Given the higher allelic fraction, c.1602del was retained and c.1604del was excluded. D). Although called as separate alleles, c.1A>C and c.2_5del lie *in cis* and represent a single multi-nucleotide variant, c.1ATGTT>C, that is also *in cis* with a 7bp deletion in the 5'-UTR.

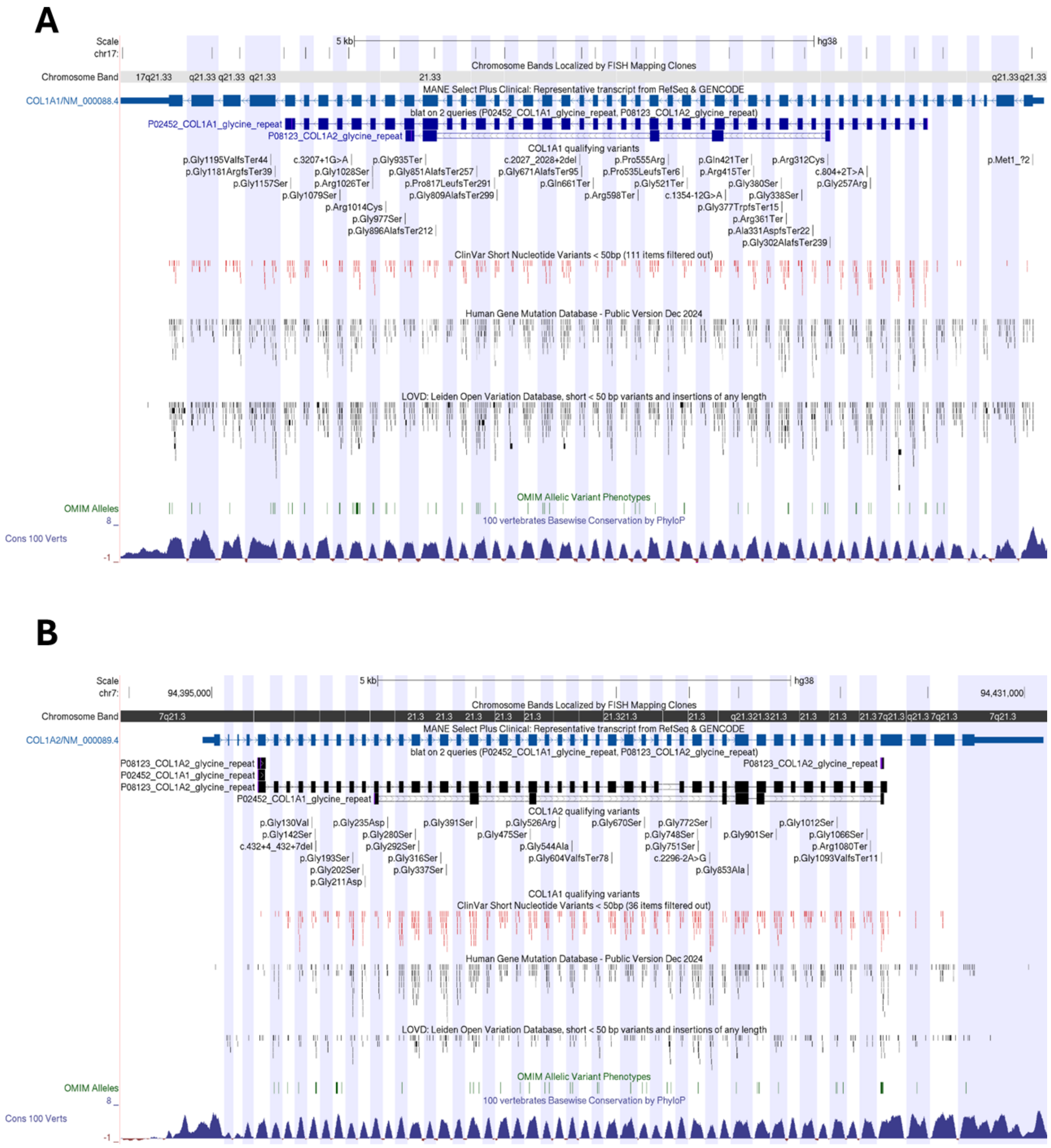


**Figure S4:** Distribution of qualifying variants in COL1A1 (A) and COL1A2 (B). For indels, genomic coordinates are left aligned and correspond to the LHS anchoring base, as per vcf standards. cDNA and protein coordinates are based on NM_000088.4 and NM_000089.4. Interactive UCSC browser sessions are available at https://genome.ucsc.edu/s/AlistairP/COL1A1_distribution and https://genome.ucsc.edu/s/AlistairP/COL1A2_distribution. Amino acid sequences in fasta format were downloaded from www.uniprot.org (accessions P02452 and P08123). All glycine-altering variants identified in this study lay within the Gly-Xaa-Yaa repeat regions for which the regions are highlighted using the BLAT search track. The ClinVar track is filtered for just missense changes that are (L)P and this shows that there is a paucity of this class of pathogenic variant before the repeat region.

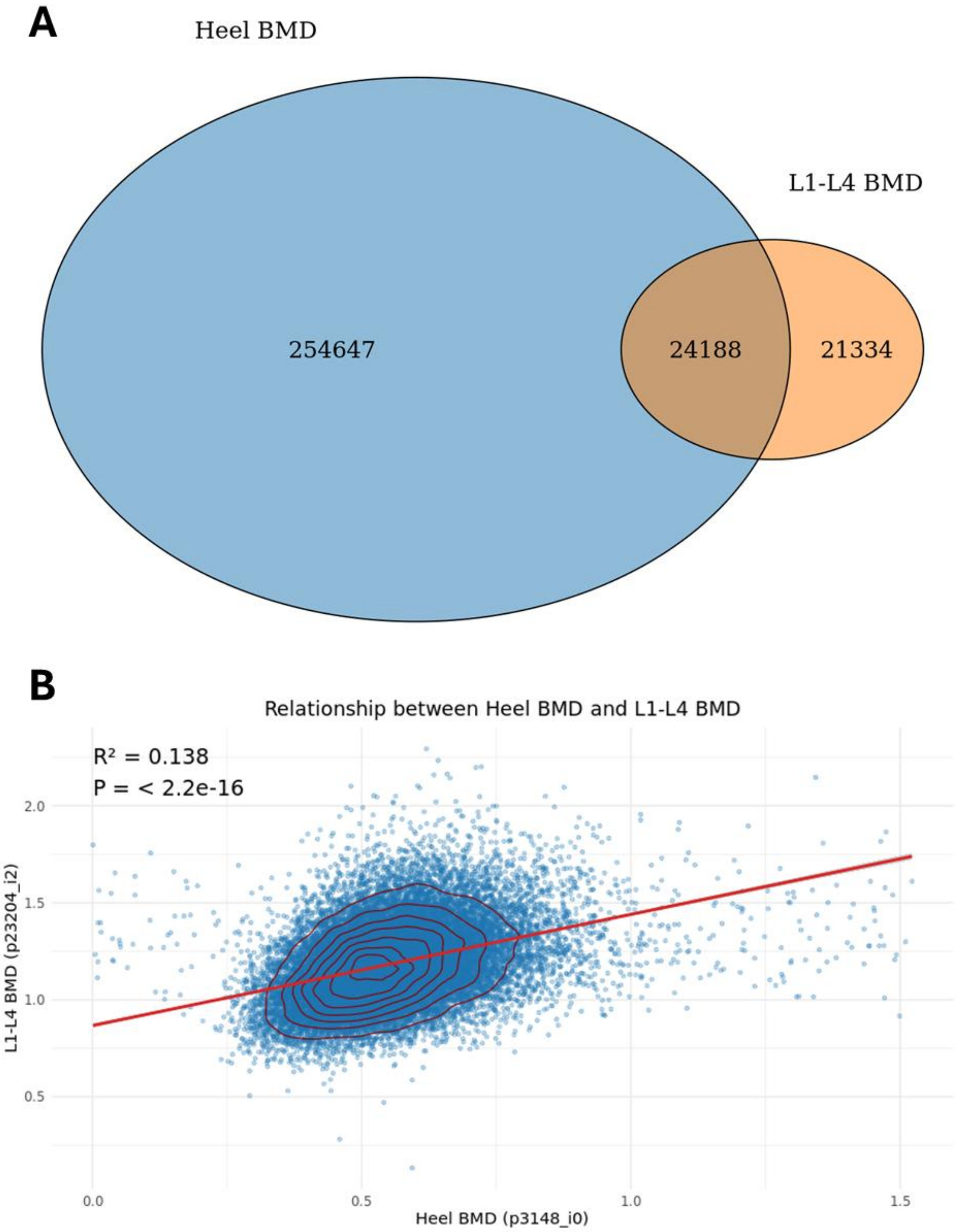


**Figure S5:** Sample overlap and correlation between heel and L1-4 Bone Mineral Density results. A) Venn diagram shows that for the 278,835 individuals with heel BMD (p3148_i0) and 45,522 with L1-L4 BMD (p23204_i2), there were 24,188 where both measurements were available. Data presented using the VennDiagram package in RStudio Pro (v2024.04.2). B) Scatter plots with linear regression trend lines and 2D density contours were generated using the ggplot2.[10] Linear regression was used to assess the association between heel and L1–L4 BMD. $R^2$ and p-values (t-test on the slope) are shown.

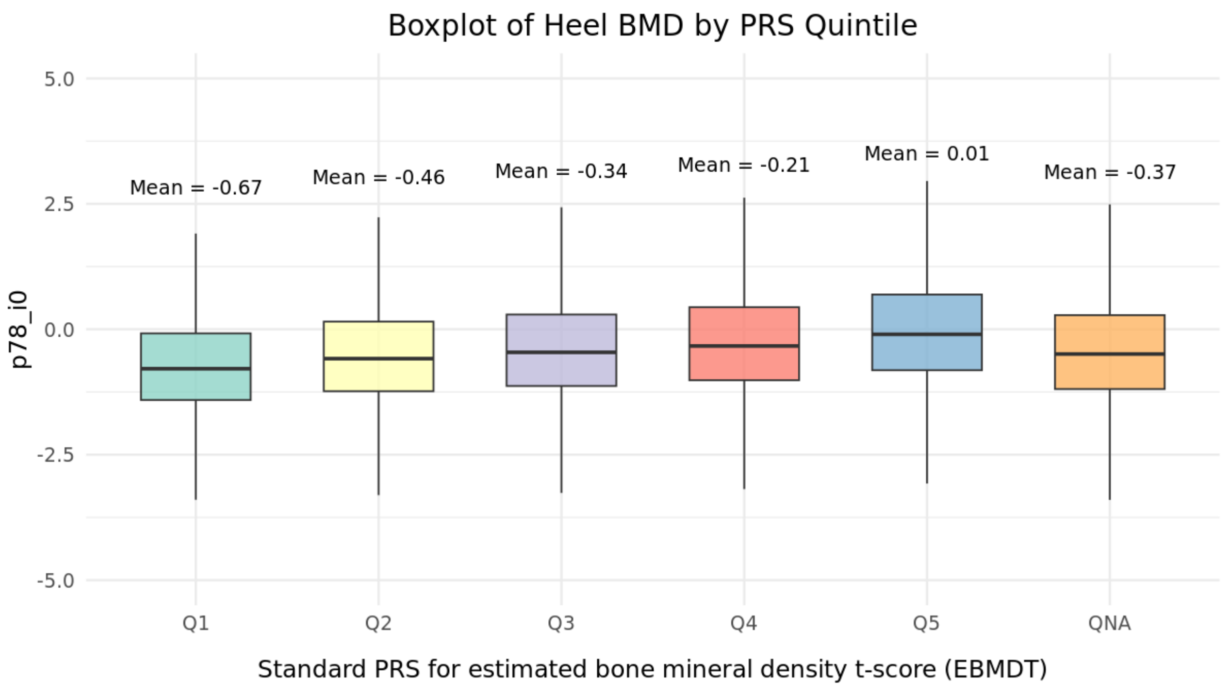


**Figure S6:** Effect of polygenic risk score (PRS) on heel bone mineral density. UK Biobank participants were divided into quintiles using the standard PRS data available in the UKB which were calculated as described previously.[11] Heel bone mineral density (p78_i0) was positively correlated with PRS scores and there was a 0.68 different between the means for quantile 1, compared to quintile 5. QNA, individuals for whom a PRS score was not available.

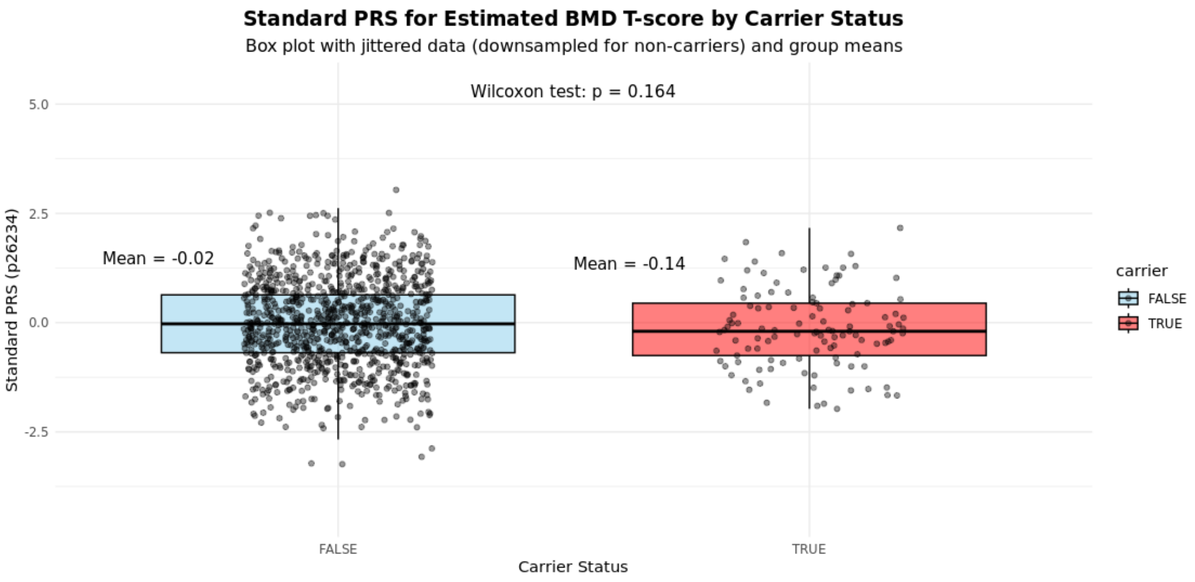


**Figure S7:** Polygenic risk scores (PRS) for Bone Mineral Density (BMD) are not increased in carriers of rare (likely) pathogenic variants in *COL1A1* or *COL1A2*. PRS data was available for 486,053 UK Biobank participants in total and for 113/115 of the rare variant carriers. Although means and statistical testing is performed using all non-carriers, for visualisation the jitter-plot is down-sampled by a factor of 0.002.
